# Assessing the barriers and enablers of on-the-job provider training for subcutaneous depot medroxyprogesterone acetate (DMPA-SC) in the public sector in Ghana: a cross-sectional mixed-methods study

**DOI:** 10.1101/2025.06.01.25328751

**Authors:** Chelsey Porter Erlank, Mensimah Bentsi-Enchill, Samuel Tagoe, Joyce Ami Amedoe, Melinda Stanley, Claudette Ahliba Diogo, Kofi Issah

## Abstract

Subcutaneous depot-medroxyprogesterone acetate is an injectable contraceptive that can be administered by any trained person, including for self-injection. When rolling out in-service provider training on subcutaneous depot-medroxyprogesterone acetate between 2019 and 2021, Ghana Health Service tasked a cohort of formally classroom-trained providers to provide on-the-job training to colleagues to cost-effectively improve training coverage. This cross-sectional mixed-methods study assessed the barriers and enablers of this approach.

The study was conducted in 2021 and included a structured quantitative survey of providers (n=192) across trained facilities in four regions in Ghana, plus key informant interviews with regional resource team members (n=8), facility in-charges (n=8), and formally-trained or on-the-job trained providers (n=16). Descriptive statistics and Chi-2 tests were used to compare quantitative outcomes between formally-trained and on-the-job trained providers in survey data. Qualitative results were analysed thematically and triangulated with quantitative results.

Most participants reported that on-the-job training had been inconsistently implemented. Where implemented, on-the-job training was reported to vary in length and depth, with on-the-job trained providers typically reporting fewer training components than formally-trained providers. Contemporary challenges around stock availability affected providers’ motivation to prioritize on-the-job training. Formally-trained providers were more satisfied with training than on-the-job trained providers (98.1% versus 50.6%, p<0.05). Both cohorts scored comparably in injectables counselling role-plays and demonstrated comparable attitudes towards injectables. However, formally-trained providers scored better in recall of the five critical self-injection steps (65.4% and 37.9%, p<0.01). Post-training supervision was highly valued but inconsistently implemented. Regional resource team members and facility in-charges recommended more supervision to standardise on-the-job training and address residual knowledge gaps.

This study found that, when inconsistently implemented, on-the-job training led to variations in provider satisfaction with training and provider skill to counsel on subcutaneous depot-medroxyprogesterone acetate, particularly for self-injection. The authors recommended standardizing on-the-job training and increasing supportive supervision to optimize the model.

## Introduction

Intramuscular depot medroxyprogesterone (DMPA-IM) is the most popular contraceptive method in Ghana representing 38.4% of the method mix. [1] In 2017, a new injectable contraceptive product - subcutaneous depot medroxyprogesterone acetate (DMPA-SC) - was registered with the Food and Drug Authority (FDA) of Ghana. With a smaller needle and all-in-one Uniject system, DMPA-SC is simpler to administer than DMPA-IM and the FDA in Ghana approved DMPA-SC with a label including indication for client self-injection (SI) as well as provider-administration (PA). DMPA-SC, including for SI, has been demonstrated as a feasible and acceptable method to clients and providers in a range of contexts, with the potential to increase continuation rates among injectable users and reduce the burden of repeat visits to facilities. [2, 3, 4, 5, 6, 7, 8, 9]

Following the registration of DMPA-SC in Ghana, the Ghana Health Service (GHS) collaborated with Population Council between 2017 and 2018 in the Ashanti and Volta regions to conduct a feasibility and acceptability study on a pilot of DMPA-SC, including for SI. [10] This included a comprehensive three-day in-service classroom-based training (50% theory, 50% practice) for the providers in the pilot. The study demonstrated high rates of feasibility and acceptability for DMPA-SC, including for the SI option, among providers and clients. [10]

Following the dissemination of the pilot study results, GHS developed a national strategic introduction and scale-up plan for DMPA-SC (both PA and SI) in 2019. [11] Under Phase 1 of the plan, several strategic objectives were achieved in 2019-2020, including integration of DMPA-SC into the in-service provider training curriculum.

Formal, in-service classroom-based provider trainings conducted between Q1 2019 and Q4 2020 trained 4,052 providers on DMPA-SC for both PA and SI, reaching 1,905 (19%) of 9,815 public and private facilities nationwide with at least one formally-trained (FT) provider by the end of 2020. The formal training consisted of a classroom component, facilitated by Regional Resource Team (RRT) members, that was similar to that conducted during the original pilot, but conducted in two days instead of three. Typically, only one provider per facility was FT in this way - except in some higher-level facilities, where two or more providers may have been selected.

Despite resource mobilization efforts, the training roll-out progress between Q1 2019 and Q4 2020 fell short of the national plan’s Phase 1 target to train 10,000 health workers across public and private sectors due to funding constraints. It has been noted that when introducing a new contraceptive method, training health workers is one of the largest cost drivers, often representing 50% or more of programme cost. [12] In an effort to expand training coverage, GHS encouraged the FT providers to share their new skills with colleagues at their facility through ‘on-the-job’ trainings (OJT). Assuming each FT provider was able to train one or two colleagues OJT, the aim of this cascaded peer-training model was to cost-effectively double or triple the coverage of DMPA-SC trained providers. However, prior to this study, it was unclear how consistently or effectively the cascaded OJT model had been implemented in practice.

Analysis conducted prior to this study indicated that although 1,905 facilities were reportedly staffed with DMPA-SC FT providers as of Q4 2020, the number of facilities reporting DMPA-SC clients (both PA and SI) into the national health information system (DHIMS) had never exceeded 900 facilities per month and averaged only 645 facilities per month in Q1 2021. Only 16% of facilities, on average, were reporting any provision of DMPA-SC for SI specifically. It was unclear if this discrepancy reflected challenges with accurate data capture for DMPA-SC or gaps in implementation and effectiveness of the OJT model.

GHS field visits across Ghana prior to this study had raised concerns that the OJT model was being implemented inconsistently in practice, leading to varied quality of client counselling on DMPA-SC, particularly for the SI option. While the rate of clients preferring the SI option can vary across contexts depending when clients are asked [4, 13], studies from other contexts have also shown that considerable time is often needed to train and reassure women about this new concept [5, 7], which can place a burden on busy providers. [14]

In 2021, as GHS and partners started to review progress under Phase 1 of the DMPA-SC roll out plan, they wanted to unpack the barriers and enablers affecting the implementation of the OJT model and how this may be impacting uptake of DMPA-SC (including for SI) in practice. Specific research questions that focused on evaluating how consistently the OJT model was being implemented and whether providers receiving the different types of training (FT versus OJT) were equally knowledgeable and confident in their skills in counselling on DMPA-SC, including for SI.

## Materials and methods

This cross-sectional observational study comprised a parallel convergent mixed-methods approach, drawing on:

1. Quantitative structured survey with providers (both FT and those eligible for OJT training – regardless of whether they had received OJT) at sampled public and private facilities
2. Qualitative key informant interviews (KIIs) with stakeholders involved in provision of in-service training, namely RRTs; facility in-charges at sites with at least one FT provider; and a sub-sample of family planning (FP) providers (both FT and OJT trained)

The primary outcome of interest was provider knowledge of the five critical steps for self-injection, as defined by GHS’ in-service training manuals. Other outcomes of interest included: perspectives on FT and OJT training received, reasons for not providing or receiving OJT, clinical knowledge around DMPA-SC, attitudes towards injectables, skill in injectables counselling and self-injection client training roleplays, and barriers and enablers of integrating DMPA-SC into routine care. Where appropriate, these outcomes were compared between untrained/self-trained, FT and OJT provider groups.

### Study sites and samples

At the time of designing the study, some of Ghana’s 16 regions had slightly higher proportionate coverage of ‘trained sites’ (i.e. sites with at least one FT provider) than others, ranging from under 5% to over 25%. The four regions for the study were purposively sampled based on being the region(s) in each Belt with the highest coverage of ‘trained sites’.

This approach was taken to ensure inclusion of facilities (and therefore providers) with sufficient exposure to the DMPA-SC in-service training to provide valuable insights. Two regions were sampled from the Southern Belt as it had the highest concentration of trained providers in the country. The sampled regions were:

- Southern Belt: Central and Eastern regions (26% and 17% coverage of trained sites at the time of the study, respectively)
- Middle Belt: Ashanti region (17% coverage)
- Northern Belt: Upper East region (9% coverage)

Within the four selected regions, districts with at least 10% training coverage (measured as the number of sites with at least one FT provider divided by total number of sites providing contraceptive services) were included in the sampling frame (n=101 out of a possible 113). Within these districts, it was estimated that 62 facilities needed to be sampled to achieve sufficient sample size of FT and OJT providers to allow for statistical testing of the primary outcome (correct clinical knowledge of DMPA-SC SI steps) between the FT and OJT populations. Sample size estimates were generated assuming: 1) an average cluster size of three providers per site, where we assumed one provider was FT and two were OJT-eligible; 2) a difference in clinical knowledge of DMPA-SC of 20 percentage points or more between FT and OJT-eligible populations; 3) intraclass correlation of 0.5; with type I error set to 1.96 and type II error set to 0.84. For the purposes of easy division for probability proportional to size (PPS) sampling, the estimated requirement of 62 facilities was increased slightly to 64 sites.

The 64 public and private ‘trained sites’ were sampled from 16 eligible districts in the four regions using PPS, such that districts with a larger population of ‘trained sites’ were more likely to be sampled. Then, within each of the 16 districts sampled, an even number of facilities (four per district) with at least one FT provider were randomly sampled. Within each site, all FT providers and OJT-eligible providers were eligible and invited to be surveyed, including those who were OJT-eligible but untrained/self-trained, as it was not possible to estimate a priori what percentage of the OJT-eligible population would have actually received OJT.

The sampling approach for the provider survey did not aim to be nationally representative. Instead, it aimed to be representative only of providers at ‘trained sites’ (i.e. with at least one FT provider) in the four purposively sampled regions. Challenges with identifying a complete sampling frame for this study are outlined in the Limitations section.

The qualitative sample was recruited from a sub-sample of the same 64 sites and spread evenly across the four regions to ensure perspectives from each region. The final qualitative sample size was based on theoretical saturation, which refers to the point at which no new concepts emerge from qualitative data drawn from a sample that is diverse in pertinent experiences and characteristics. [15] In order to assess whether theoretical saturation had been achieved, transcription was conducted in an iterative way throughout the data collection process and initial transcripts reviewed by the coding team. Table 1 outlines the final quantitative and qualitative sample sizes.

**Table 1.**
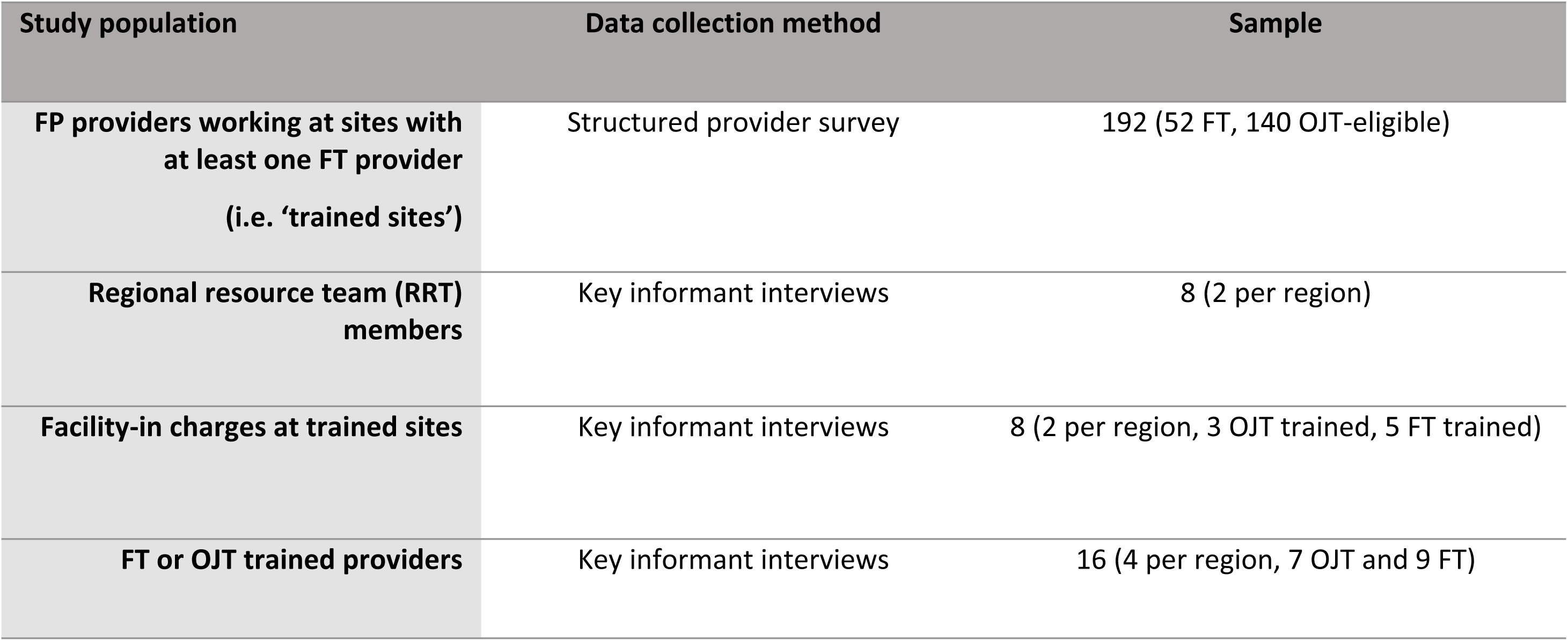
Sample size by population and data collection method.

### Data collection

Quantitative data collection tools for the provider survey were developed to align with content of the GHS’ in-service training curriculum on DMPA-SC. The survey included questions on experience of FT or OJT training (if received); a counselling role-play where the data collector played the role of a ‘client’ interested in injectables and scored the providers according to set criteria from GHS counselling guidelines; a client-training role-play where the data collector played the role of a ‘client’ interested in learning to SI and scored the providers on their recall of the five critical SI steps as defined in GHS training manuals (1: select appropriate injection site and clean it if needed; 2: Mix the solution by shaking the device for about 30 seconds; 3: Activate the device by closing the gap between the needle cap and port; 4: Gently pinch the skin at the injection site to form a ‘tent’; 5: Insert needle completely so that port touches skin and press the reservoir slowly to inject for about 5 to 7 seconds); Likert scale questions assessing provider attitudes towards injectables; and a true/false test of knowledge about DMPA-SC. Qualitative KII discussion guides were semi-structured and developed based on a theoretical framework mapping out a provider behaviour change journey towards integration of DMPA-SC, including for SI, into routine care. This framework was developed from a rapid literature review conducted prior to the study and is outlined in S1 Table.

Qualitative interviewers and quantitative data collectors participated in a three-day training workshop, including training on how to assess injectables counselling and SI training against the assessment criteria, as well as an opportunity to pre-test and revise the study tools. Subsequently, participants were recruited and surveyed and/or interviewed over a two-week period (1^st^ to 12^th^ November) in 2021. Site substitutions from the same district were necessary in cases where the originally sampled sites did not have an FT provider in post at the time of the study (for example, due to incorrect data from original FT trainings, attrition or leave) – see the Limitations section of this paper for more details. Response rates were high, with very few cases of provider refusal to participate. Quantitative data was collected through tablets into open-source software (Kobo Collect), while qualitative data was audio-recorded and transcribed verbatim in the language of interview (mostly English, although some were conducted in Akan or Frafra before being translated into English for analysis).

### Data analysis

Quantitative data was exported from Kobo Collect, cleaned and analysed as descriptive statistics initially in SPSS by one member of the analysis team (ST). Differences between the FT and OJT sub-groups in the provider survey were tested using the SVY syntax (to account for clustering by facility) and Chi2 tests in STATA-17 by another member of the team (CPE). Missing data was minimal in the quantitative survey dataset, limited to some of the variables around training length and provision of OJT. There was no missing data in any of the outcome variables being statistically tested. Where all providers had responded to a specific question, initial Chi2 tests were run comparing the three groups (untrained/self-trained, FT, and OJT), before being re-run just between FT and OJT groups to assess the significance of any differences. All quantitative findings were then triangulated with qualitative findings by four members of the team (CPE, ST, MS, MBE) for contextualization, in line with a convergent mixed-methods approach.

Three members of the study team (CPE, MS and ST) coded 25% of the qualitative transcripts for each of the three KII populations deductively at first against a draft set of codes developed based on the provider-side theoretical framework in S1 Table. These independent code applications were then compared to check for consistency and additional inductive codes added to create a comprehensive codebook. Once consistency in code application was achieved between the coding team, the final codebook was then applied to all remaining transcripts by the same team of coders. Dedoose software was used for data coding, organization and retrieval.

Validation workshops with key stakeholders in the study (RRT members, Public Health Nurses, selected facility in-charges, and selected FT and OJT providers) were conducted in two study regions (Ashanti and Eastern) in June 2022 to share the draft findings, check interpretation, and help with generating recommendations.

### Ethics statement

This study was conducted in line with the principles outlined in the Declaration of Helsinki. The study team worked closely with the study regions, districts and facilities through the office of the GHS Director General to secure appropriate approvals. The study protocol and tools were approved by GHS Ethics Review Committee in Accra, Ghana, a board that is also registered as an International Institutional Review Board with Federal Wide Assurance (IRB: IRB00009260, FWA00020025), study protocol approval number GHS-ERC: 019/08/21. All participants provided written informed consent to participate in the survey and KIIs.

## Results

### Participant characteristics

The structured provider survey included a cadre mix of FP providers and facility in-charges (n=192) from the 64 sampled facilities - the majority (69.3%) were community health nurses (CHNs), while 16.7% were nurses, 10.4% were midwives and 3.6% had other cadres (for example, field technicians or health assistants). Facility-in charges and providers (n=24) participating in the KIIs included a mixture of CHNs, midwives, and other nurses. They reported working at a range of public and private sites - mainly community-based health planning and service (CHPS) facilities or health centres and hospitals in a few cases. All sampled RRTs (n=8) were involved in the facilitation of the DMPA-SC in-service formal training in their region; some had additionally been involved in routine supportive supervision visits since the trainings.

### Implementation of the OJT model in practice

In the survey, nearly three quarters (72.4%) of providers had received some DMPA-SC training (27.1% FT plus a further 45.3% OJT) while the remaining 27.6% had not been trained yet, despite working at a site with at least one FT colleague (Table 2). One provider surveyed did not receive any training but was self-taught via the internet. This provider was grouped with the untrained group for further analysis. For every one FT provider in the survey, on average another 2.7 FP providers at the same sites were eligible for OJT, but in practice, only approximately 1.7 had received the OJT at the time of the survey - despite most FT providers having received their FT between seven and twelve months prior to the survey (Table 2). Both FT providers and untrained providers in the survey were asked to indicate the main reasons for not yet providing or receiving OJT - most agreeing that colleague unavailability and insufficient tools were the main reasons (Table 2).

**Table 2.**
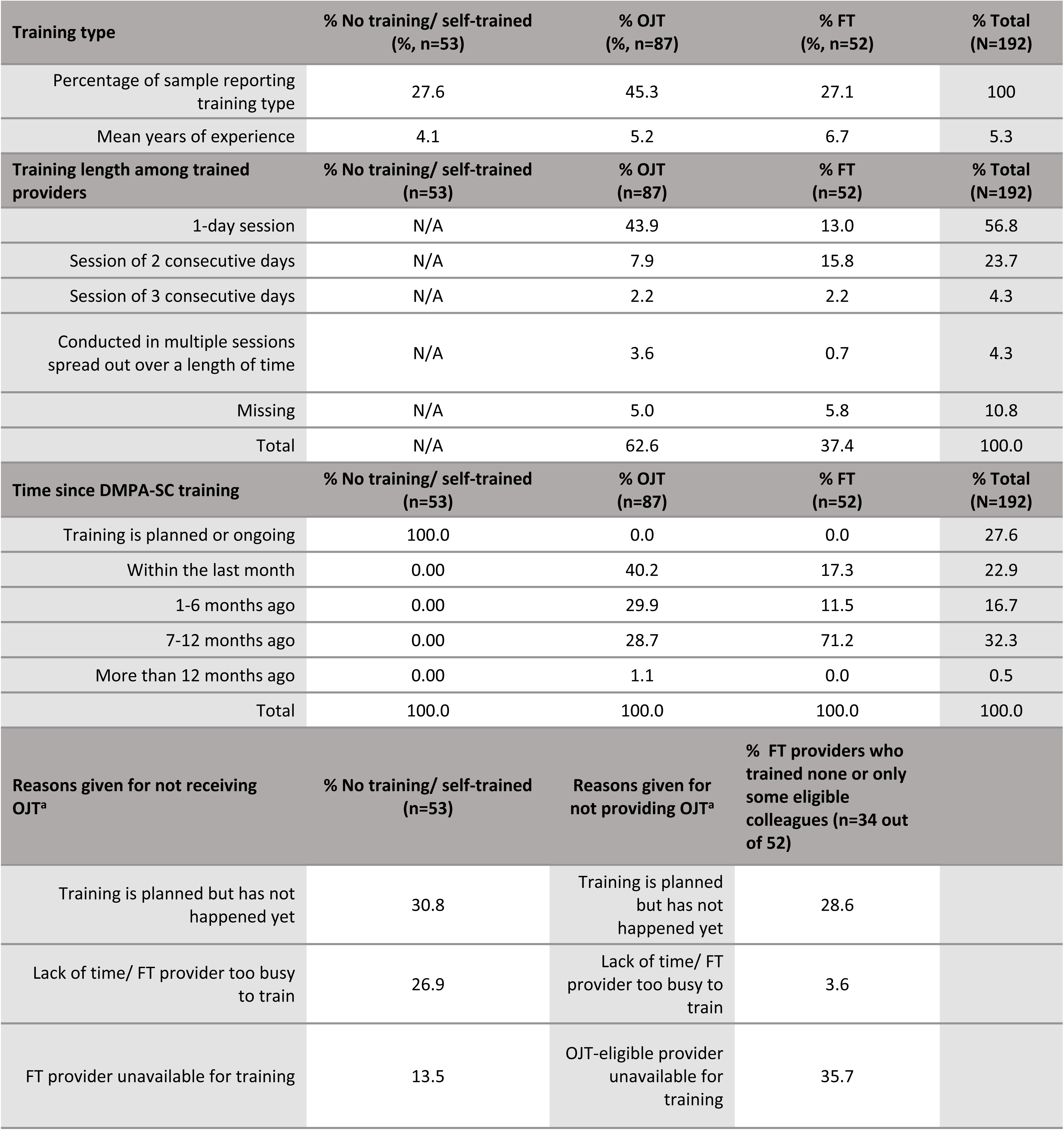
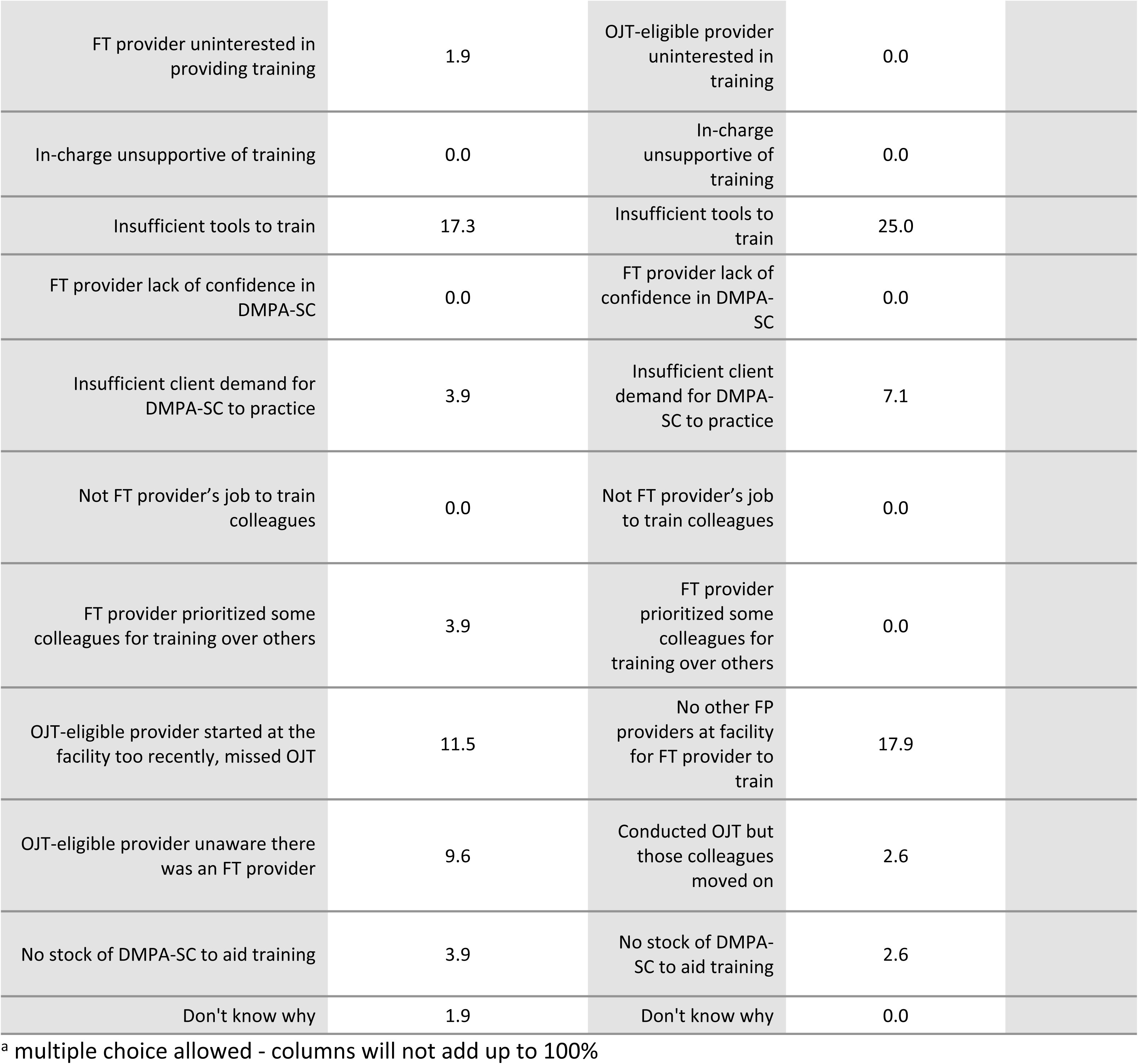
Survey responses about training provided and/or received on DMPA-SC.

| Training type | % No training/ self-trained<br>(%, n=53) | % OJT<br>(%, n=87) | % FT<br>(%, n=52) | % Total<br>(N=192) |
| --- | --- | --- | --- | --- |
| Percentage of sample reporting training type | 27.6 | 45.3 | 27.1 | 100 |
| Mean years of experience | 4.1 | 5.2 | 6.7 | 5.3 |
| Training length among trained providers | % No training/ self-trained<br>(n=53) | % OJT<br>(n=87) | % FT<br>(n=52) | % Total<br>(N=192) |
| 1-day session | N/A | 43.9 | 13.0 | 56.8 |
| Session of 2 consecutive days | N/A | 7.9 | 15.8 | 23.7 |
| Session of 3 consecutive days | N/A | 2.2 | 2.2 | 4.3 |
| Conducted in multiple sessions spread out over a length of time | N/A | 3.6 | 0.7 | 4.3 |
| Missing | N/A | 5.0 | 5.8 | 10.8 |
| Total | N/A | 62.6 | 37.4 | 100.0 |
| Time since DMPA-SC training | % No training/ self-trained<br>(n=53) | % OJT<br>(n=87) | % FT<br>(n=52) | % Total<br>(N=192) |
| Training is planned or ongoing | 100.0 | 0.0 | 0.0 | 27.6 |
| Within the last month | 0.00 | 40.2 | 17.3 | 22.9 |
| 1-6 months ago | 0.00 | 29.9 | 11.5 | 16.7 |
| 7-12 months ago | 0.00 | 28.7 | 71.2 | 32.3 |
| More than 12 months ago | 0.00 | 1.1 | 0.0 | 0.5 |
| Total | 100.0 | 100.0 | 100.0 | 100.0 |
| Reasons given for not receiving OJT<sup>a</sup> | % No training/ self-trained<br>(n=53) | Reasons given for not providing OJT<sup>a</sup> | % FT providers who trained none or only some eligible colleagues (n=34 out of 52) |  |
| Training is planned but has not happened yet | 30.8 | Training is planned but has not happened yet | 28.6 |  |
| Lack of time/ FT provider too busy to train | 26.9 | Lack of time/ FT provider too busy to train | 3.6 |  |
| FT provider unavailable for training | 13.5 | OJT-eligible provider unavailable for training | 35.7 |  |
| FT provider uninterested in providing training | 1.9 | OJT-eligible provider uninterested in training | 0.0 |  |
| In-charge unsupportive of training | 0.0 | In-charge unsupportive of training | 0.0 |  |
| Insufficient tools to train | 17.3 | Insufficient tools to train | 25.0 |  |
| FT provider lack of confidence in DMPA-SC | 0.0 | FT provider lack of confidence in DMPA-SC | 0.0 |  |
| Insufficient client demand for DMPA-SC to practice | 3.9 | Insufficient client demand for DMPA-SC to practice | 7.1 |  |
| Not FT provider's job to train colleagues | 0.0 | Not FT provider's job to train colleagues | 0.0 |  |
| FT provider prioritized some colleagues for training over others | 3.9 | FT provider prioritized some colleagues for training over others | 0.0 |  |
| OJT-eligible provider started at the facility too recently, missed OJT | 11.5 | No other FP providers at facility for FT provider to train | 17.9 |  |
| OJT-eligible provider unaware there was an FT provider | 9.6 | Conducted OJT but those colleagues moved on | 2.6 |  |
| No stock of DMPA-SC to aid training | 3.9 | No stock of DMPA-SC to aid training | 2.6 |  |
| Don't know why | 1.9 | Don't know why | 0.0 |  |

<sup>a</sup> multiple choice allowed - columns will not add up to 100%

In KIIs, FT providers’ reported experience of implementing the OJT varied, with a few FT providers reporting actively running regular (for example, twice per year) trainings for their colleagues, while the majority reported holding only one initial post-training ‘briefing’ for their colleagues. Most OJT providers in the survey reported a one-day training (Table 2) which they clarified in KIIs was typically just a single short session or ‘briefing’ on DMPA-SC, with the chance to ask questions and perhaps a demonstration with one Uniject, where samples were available.

FT providers answering the survey were more likely to report receiving a wide range of elements of the training compared to OJT providers (Fig 1). All differences in Fig 1 were statistically significant (p<0.01) except for ‘e) one-to-one supportive supervision’ (p=0.34) and ‘g) the opportunity to discuss/ask questions’ (p=0.07).

**Fig 1.**
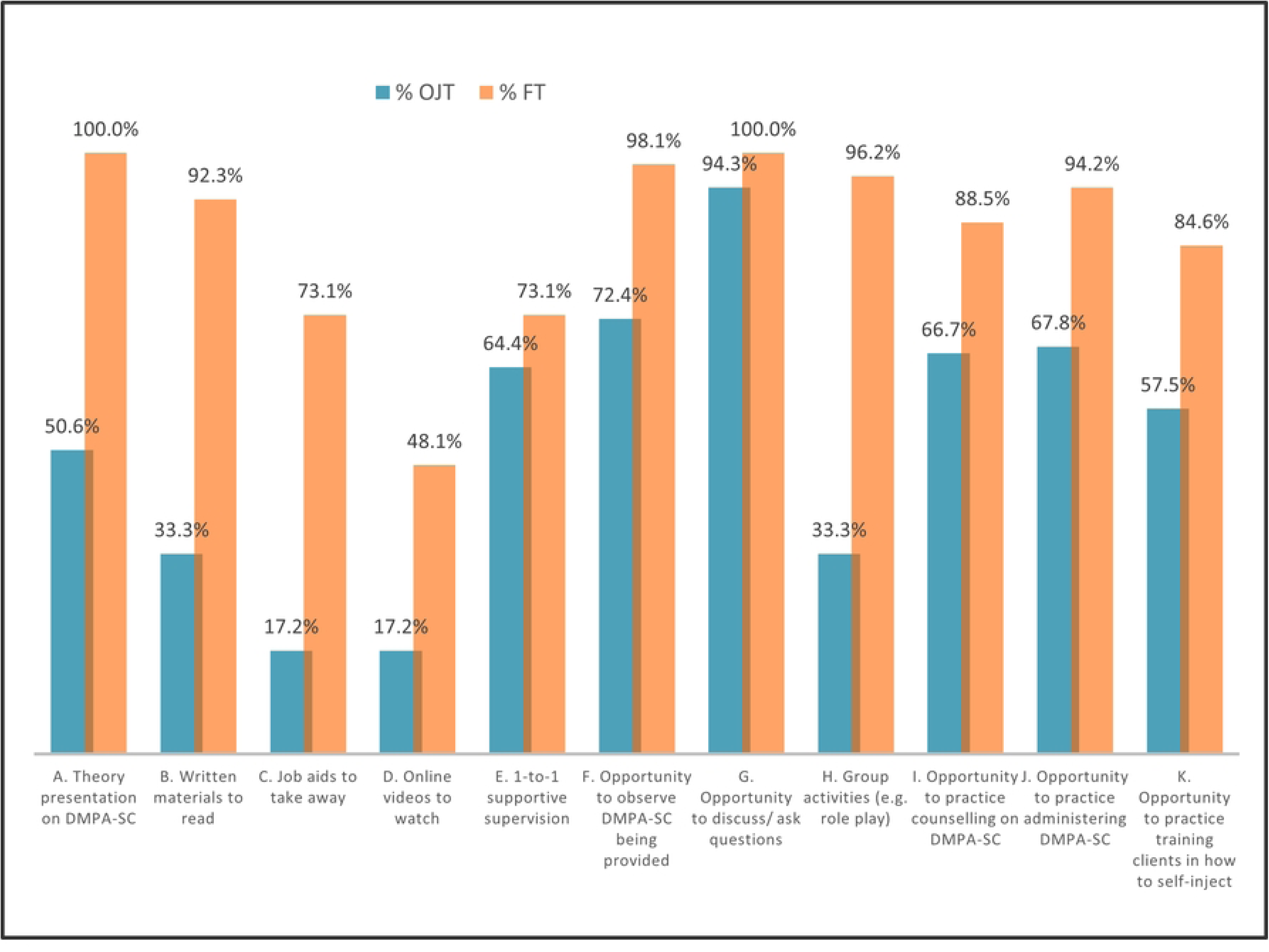
Training components by training type (OJT vs FT)

Just over half (56.8%) of providers/in-charges said that their facility was providing DMPA-SC at the time of the survey, while 30.7% said it was not, and 12.5% said they did not know. In the survey, 40.4% of providers who were aware of DMPA-SC being provided at their facility (n=109) said they had faced challenges with injectables stockouts in the last three months (68.2% of these said the issue was with DMPA-SC, while 15.9% said DMPA-IM and 15.9% said both injectable products). In the KIIs, some providers reported not receiving any DMPA-SC commodities at all since being trained. Others only received limited commodities that were meant to be used for demonstration during OJT, while a few said initial stocks for service provision ran out after a few months. Aside from disrupting provision of DMPA-SC, some providers also talked about stockouts having disruptive effects on FT providers’ ability to train their colleagues OJT:

> *“I have been able to talk to my colleagues [provide OJT training], but they have not seen the commodity before, I have explained things to them that this is how it is, this is how it’s been used and the side effects and other things, but they have not yet seen the commodity nor practiced with it.” - **FT provider, Ashanti***

### Providers’ satisfaction with training received

In the KIIs, all RRTs interviewed said that by the end of the DMPA-SC trainings they were confident the FT providers had grasped the method. Most RRTs and FT providers in the KIIs felt that the two-day length of the in-service FT was sufficient, because the participants were already experienced FP providers, while a minority of both groups felt that the trainings should be extended by another day to ensure sufficient time for practice.

Over two thirds (68.4%) of providers in the survey reported overall satisfaction with their training, however this differed significantly between FT and OJT providers (98.1% versus 50.6% respectively, p<0.01). In fact, OJT providers were statistically significantly less satisfied with almost every element of their training (p<0.05) except post-training support (where they were slightly more satisfied, p=0.04) and venue (where the difference did not emerge as significant, p=0.25) (Fig 2).

**Fig 2.**
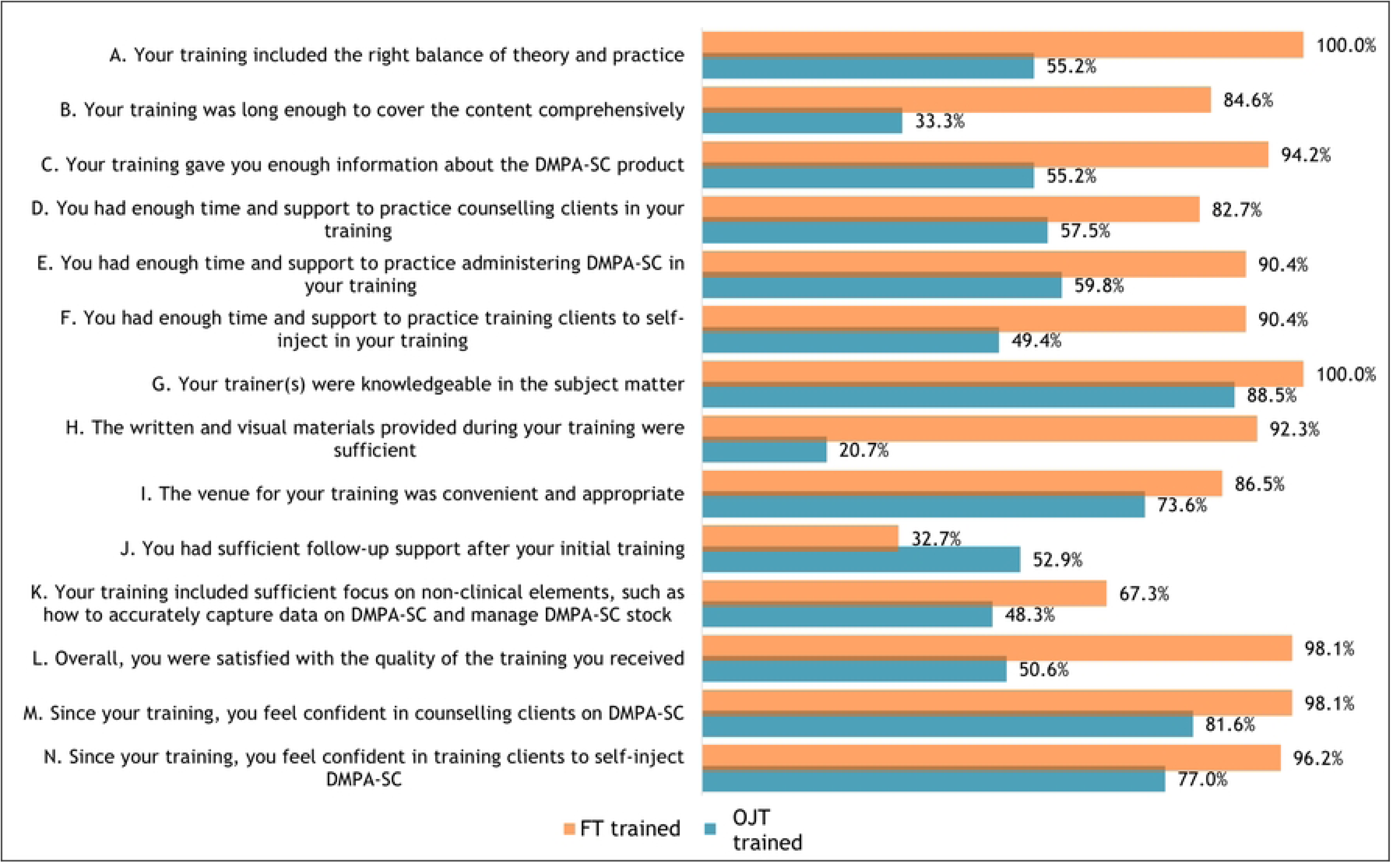
Percentage respondents agreeing/strongly agreeing to statements on quality of training received in DMPA-SC by training type (OJT vs FT)

In KIIs, most FT providers were positive about the content of their training - they particularly appreciated the visual/written aids they were given, seeing the Unijects, and the opportunity for discussion and role plays with colleagues.

> *“…the training materials were good, they showed us how the Sayana Press [DMPA-SC] is used, they even opened it and we did practical on it and the trainers were good.” – **FT provider, Upper East***

For their part, while seemingly hesitant to criticize their FT colleagues, several OJT providers in KIIs indicated that the insufficient level of the information received and lack of stock (in some cases) to facilitate demonstrations and practice opportunities meant their DMPA-SC training was insufficient.

> *“… when they [FT colleagues] came to train us, we used one [DMPA-SC unit] for the training for example, but we didn’t inject on anybody so as to see how it is opened and how to even pierce [activate] it…” - **OJT provider, Eastern***

### Providers’ post-training attitudes, knowledge and skills on provision of DMPA- SC

#### Provider attitudes towards injectables post-training

The provider survey included a series of attitudinal statements about injectable contraceptives and providers were asked if they agreed or disagreed with the statements. While significant differences in attitudes were found between trained and untrained providers on some statements (p<0.05 for statements b, d, f), there were no significant differences between FT and OJT providers, except that FT providers were more likely than OJT providers to agree/strongly agree they have ‘no time to train women in SI on busy days’ (p=0.04) – possibly reflecting these providers’ more senior status within the facilities (Fig 3).

**Fig 3.**
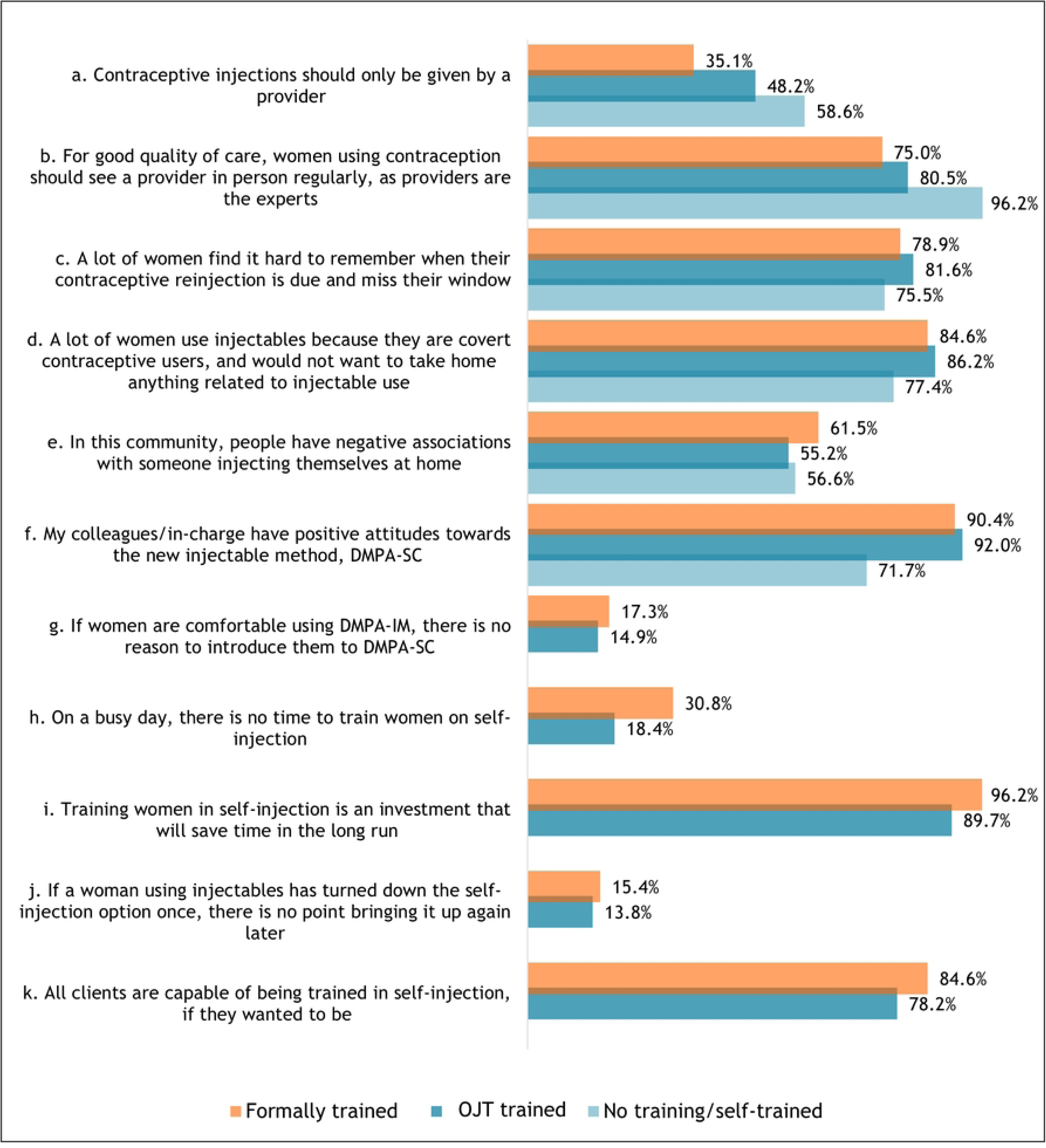
Percentage of providers who agree / strongly agree with attitudes statement, by training type

#### Post-training knowledge retention on DMPA-SC

In the survey, providers with any training (FT or OJT) showed knowledge of DMPA-SC that was statistically significantly better than the untrained group - who unsurprisingly tended to respond that they did not know the answer in most cases (all p<0.01, Table 3). When omitting the untrained/self-trained group, FT providers were statistically significantly more likely to score >85% correct answers than their OJT counterparts (59.6% FT versus 26.4% OJT, p<0.01), driven mainly by significant differences in knowledge on dosage of DMPA-SC and correct disposal routes for used Unijects (both differences p<0.01).

**Table 3.** Survey results for post-training knowledge assessment.

| DMPA-SC knowledge statements (N=192) | Response (correct response highlighted in green) | No training/self-trained (n=53) | OJT trained (n=87) | Formally trained (n=52) |
| --- | --- | --- | --- | --- |
| a. DMPA-SC (Sayana Press) does not need to be activated before use | TRUE | 9.40% | 19.50% | 19.20% |
|  | FALSE | 11.30% | 64.40% | 76.90% |
|  | I don't know | 79.30% | 16.10% | 3.90% |
| b. DMPA-SC (Sayana Press) contains a lower dosage of active hormone compared to DMPA-IM | TRUE | 11.30% | 35.60% | 59.60% |
|  | FALSE | 15.10% | 35.60% | 32.70% |
|  | I don't know | 73.60% | 28.70% | 7.70% |
| c. Used units of DMPA-SC can be disposed of in latrines | TRUE | 9.40% | 33.30% | 13.50% |
|  | FALSE | 35.90% | 58.60% | 84.60% |
|  | I don't know | 54.70% | 8.10% | 1.90% |
| d. DMPA-SC differs from DMPA-IM because it can be injected into the skin, rather than the muscle | TRUE | 22.60% | 63.20% | 69.20% |
|  | FALSE | 9.40% | 28.70% | 28.90% |
|  | I don't know | 67.90% | 8.10% | 1.90% |
| e. DMPA-SC should not be used by women who are breastfeeding | TRUE | 7.70% | 6.90% | 9.80% |
|  | FALSE | 36.50% | 82.80% | 86.30% |
|  | I don't know | 55.80% | 10.30% | 3.90% |
| f. Clients who received DMPA-IM three months ago cannot switch to DMPA-SC during their reinjection window | TRUE | 3.80% | 11.50% | 9.60% |
|  | FALSE | 34.00% | 82.80% | 90.40% |
|  | I don't know | 62.30% | 5.80% | 0.00% |
| g. DMPA-SC protects against pregnancy for 1 month only | TRUE | 0.00% | 2.30% | 3.90% |
|  | FALSE | 43.40% | 95.40% | 92.30% |
|  | I don't know | 56.60% | 2.30% | 3.90% |

#### Assessment of role-played injectables counselling

In the survey, all providers were asked to role play a counselling session with a ‘client’ (the data collector) who was only interested in injectables. They were asked to use all the messages and props they would typically use with a real client. Data collectors were trained to score the providers’ counselling approach against a set of criteria (drawn from the national DMPA-SC training curriculum). Generally both FT and OJT providers scored better than untrained providers – especially in terms of mentioning the SI option and offering to train the client in SI or refer them for SI training (both differences p<0.01). While some minor differences emerged in the responses between the FT and OJT groups, none were large enough to be detected as statistically significant (all p>0.05, Table 4).

**Table 4.** Survey results for assessment of injectable counselling and SI training.

| Data collector assessment of injectable counselling (N=192) | Assessment outcome | No training/self-trained (n=53) | OJT trained (n=87) | Formally trained (n=52) |
| --- | --- | --- | --- | --- |
| Provider asked appropriate questions about fertility intentions and lifestyle | Yes | 43.4% | 59.8% | 61.5% |
|  | Partially | 18.9% | 21.8% | 19.2% |
|  | No | 37.7% | 18.4% | 19.2% |
| Provider presented all injectable options equally and did not try to convince use of one method over another | Yes | 45.3% | 78.2% | 88.5% |
|  | Partially | 28.3% | 13.8% | 1.9% |
|  | No | 26.4% | 8.1% | 9.6% |
| Provider explained differences between the injectable options clearly | Yes | 30.2% | 65.5% | 80.8% |
|  | Partially | 41.5% | 26.4% | 13.5% |
|  | No | 28.3% | 8.1% | 5.8% |
| Provider clearly explained the pros and cons of each injectable option | Yes | 47.2% | 63.2% | 65.4% |
|  | Partially | 22.6% | 20.7% | 21.2% |
|  | No | 30.2% | 16.1% | 13.5% |
| Provider mentioned all three injectable options (without prompting) | Yes, all three | 22.6% | 64.4% | 73.1% |
|  | Mentioned one or both PA options but not SI | 49.1% | 27.6% | 25.0% |
|  | Did not specify that there are difference types of injectable | 28.3% | 8.1% | 1.9% |

| After 'client' revealed interest in SI, provider offered to take time to train 'client' in SI OR refer 'client' to a colleague for SI training | Yes, offered to train 'client' in SI | 13.2% | 82.8% | 84.6% |
| --- | --- | --- | --- | --- |
|  | Yes, offered to refer 'client' for SI training | 30.2% | 6.9% | 5.8% |
|  | No, did not offer to train nor refer 'client' for SI training | 56.6% | 10.3% | 9.6% |
| Data collector assessment of self-injection training (n=139) | Assessment outcome | No training/self-trained (n=53) | OJT trained (n=87) | Formally trained (n=52) |
| Provider reassured and build 'client' confidence in being able to self-inject | Yes | N/A | 56.3% | 71.2% |
|  | Partially | N/A | 31.0% | 25.0% |
|  | No | N/A | 12.6% | 3.8% |
| Provider mentioned all 5 critical SI steps<sup>a</sup> | At least 1 critical step missed | N/A | 62.1% | 34.6% |
|  | All 5 critical steps mentioned | N/A | 37.9% | 65.4% |
| Provider used client SI instruction guidelines | Yes | N/A | 37.9% | 44.2% |
|  | Partially | N/A | 10.3% | 19.2% |
|  | No | N/A | 51.7% | 36.5% |
| Provider used example reinjection calendar | Yes | N/A | 31.0% | 42.3% |
|  | Partially | N/A | 18.4% | 9.6% |
|  | No | N/A | 50.6% | 48.1% |
| Provider used example disposal container | Yes | N/A | 37.9% | 65.4% |
|  | Partially | N/A | 17.2% | 7.7% |
|  | No | N/A | 44.8% | 26.9% |
| Provider used models for SI practice (e.g. condom filled with salt) | Yes | N/A | 12.6% | 34.6% |
|  | Partially | N/A | 9.2% | 7.7% |
|  | No | N/A | 78.2% | 57.7% |
| Provider used example Uniject | Yes | N/A | 60.9% | 69.2% |
|  | Partially | N/A | 12.6% | 7.7% |
|  | No | N/A | 26.4% | 23.1% |

<sup>a</sup>1. Critical step 1: select appropriate injection site / clean if needed; 2. Critical step 2: Mix solution by shaking device for about 30 seconds; 3. Critical step 3: Activate device by closing gap between needle cap and port; 4. Critical step 4: Gently pinch skin at injection site to form a 'tent'; 5. Critical step 5: Insert needle completely so that port touches skin and press the reservoir slowly for about 5 to 7 seconds

When probed in KIIs about whether they emphasized or omitted the SI option for particular types of clients, several providers did admit to using their own observations and judgments about which clients might be most interested in, or capable of, self-injecting. For some of these providers, this was more about assessing which clients might be sufficiently motivated by the benefits of SI to overcome fears (for example, those who lived far away, or travelled often). However, among a minority of providers, this judgment call seemed to reflect providers’ lack of trust that women with low levels of literacy would be able to SI.

> *“…we [providers] had some concerns because majority of our clients are also not literate so and most of the time some may even miss their [reinjection] dates … That was our major concern… How to even teach them to understand the procedure… Because you take the person through the training, and you say ‘oh do it let me see’ and then the person is all over the place.” - **FT facility in-charge, Eastern***

#### Assessment of role-played self-injection client training

After role-playing an injectable counselling session, providers in the survey were then asked to assume the ‘client’ (the enumerator) was interested in the SI option and proceed to SI training. Again, enumerators were trained to assess the providers’ approaches against set criteria, drawn from the national DMPA-SC training curriculum. Overall, 48.2% of trained providers mentioned all five critical SI steps during the role play. However, this was statistically significantly different between FT and OJT providers (65.4% and 37.9% mentioning all five critical steps respectively, p<0.01). In addition, OJT providers were significantly less likely to use an example of disposal container and demonstration materials than FT providers (both differences p<0.01). There were no other statistically significant differences detected in the SI training indicators from Table 4.

In KIIs, most providers interviewed (both FT and OJT) reported struggling with introducing the SI concept to women and reassuring them enough to SI. A small number of providers (FT and OJT) reported improvising techniques to support women’s confidence with SI, such as showing women how small the Uniject is, using visual aids or gestures, conducting repeated demonstrations, and leveraging the testimonies of experienced SI clients. While most providers in KIIs were positive about the idea of visual aids to help clients understand and recall the SI steps, many did not appear to have access to any visual aids - if they did, these were mainly one or two items left over from their own formal trainings.

Only a few (mostly FT) providers mentioned that they regularly include SI demonstrations and/or practice opportunities for women, such as using condoms filled with salt or improvised materials (e.g. oranges), in their training approach. By contrast, the majority of providers in KIIs either did not mention conducting demonstrations/practice sessions on inanimate materials or said they preferred not to conduct them, seemingly worrying about ‘wasting’ DMPA-SC stock on demonstrations and practice sessions. Instead, some providers talked about demonstrating how Unijects worked through PA on a real client and/or asking women to directly SI under observation without prior practice on inanimate objects.

### Reflections on the OJT model and the role of supportive supervision

The majority of RRTs felt that the post-training supportive supervision for the DMPA-SC in-service training roll-out had been inadequate and a few attributed this to lack of funding. Two RRT members noted that this follow-up aspect was particularly important in the OJT model, to quality assure the level of OJT provided by FT providers and correct any misunderstandings.

> *“They should be a follow-up, okay, they should be a follow up, it’s not necessary trying to bring all of them [providers] to a place to train but you train few and there should be a follow up that, you go for supervision … [of] those who were trained, you can re-access them and you get the chance also to train the new ones and you also find out that when they came [back], were they able to train others [OJT], so that you know their weakness and then you build it up, they should be frequent supervision…” – **RRT Upper East***

On their side, FT and OJT providers had mixed experiences of receiving follow-up supportive supervision (sometimes described instead as mentorship or monitoring visits) after their trainings. Around half reported receiving supportive supervision visits from a variety of sources - some from GHS, others from non-governmental organisations. The other half of FT and OJT providers said they had not received any follow-up support, except some ad hoc advice from peers or facility in-charges.

Even among those who received a post-training visit, they often expressed the need for more frequent visits. In particular, those requesting more supportive supervision wanted more support with how to introduce DMPA-SC to women in a way that reassures them about the SI option.

> *“… the supervisors … they have to come and visit us, to visit the communities to talk about the commodity… The supervisors’ have to come along with us when we go for outreach like this… so they can also talk to them [clients] for us.” - **OJT provider, Central***

All participants (RRTs, facility in-charges and providers) were asked to reflect on the effectiveness and functionality of the OJT model. The majority of RRT members were positive about the cost-effectiveness of the model, but some worried about the quality of information reaching OJT providers. The majority of FT providers were also worried about the quality of OJT they were passing on to their colleagues.

> *“[OJT model] it’s good because it will save money… I mean [in terms of] getting to train everybody, it will save time and money and then … the negative aspect of it will be that probably the person who was present at the workshop may not be able to … relay the information and be able to train the others quite properly. That may be a disadvantage.” **– FT facility in-charge, Eastern***

Most OJT providers also expressed concern that the information they had received was incomplete.

> *“I think we were trained on the job [with] this thing [DMPA-SC] … Maybe what our [FT] in-charge taught us was… she can omit something and so when the opportunity comes, [maybe] you can train some of the [other] staffs…” **- OJT provider, Ashanti***

By contrast, a few FT providers and one OJT provider described examples of the OJT model working well in their facility. These examples seemed to be reflective of the minority of cases where an experienced trainer was selected as the FT provider, and where these FT providers were running regular (for example, twice per year) and practice-based OJT:

> *“So it’s like twice a year, we go through that …like I told you, we put whatever we learnt into practice. And so since they came, they’ve been practicing and like I said, they they’ve been giving … they [FT providers] are also training their… their colleagues. So it’s like they are flowing together. We haven’t had any challenge…” - **FT facility in-charge, Central***

In fact, one FT provider said the OJT model had the benefit of there being real clients to work with, and continuous practice opportunities, as long as there was stock available.

## Discussion

This study explored the implementation of a cascaded OJT model for DMPA-SC across four regions in Ghana. The results indicated that FT providers were broadly satisfied with the breadth and quality of their two-day formal classroom training, although a few indicated that additional time to practice and written and visual materials would have improved their experience. However, the ratio of FT providers to OJT and untrained providers in the survey indicates that the OJT model was not reaching its full potential at the time of the study. In addition, the study found opportunities within the implementation of the OJT model where quality and timeliness of OJT could be strengthened.

Most FT providers (but not all) appear to have provided some form of OJT to their eligible colleagues upon their return to the facility, however this varied in timeliness, breadth and quality. Where OJT was not being provided at all, this mainly appeared to be due to lack of stock/tools to train, provider attrition or leave, and busy provider schedules. Where OJT was being provided, in most cases it seemed to consist of a short, one-off post-training debrief session by the FT provider, often without visual or written materials and sometimes also without DMPA-SC stock. It’s likely that the quality of OJT was further affected by delays in provision of OJT by FT providers.

Stock issues were a major challenge at the time of the study that had knock-on effects on all areas of integration of DMPA-SC, including the prioritisation of OJT and provider confidence in the product. In 2020, a global disruption to the manufacturing process of the sole supplier led to limited commodity availability, which constrained the scale-up and broader distribution of DMPA-SC. A manufacturing backlog, high demand worldwide, and product registration requirements in Ghana prevented the timely shipment of additional orders of DMPA-SC in the latter half of 2020, leading to facility-level stock outs and undermining the scale-up momentum – including disruption of supplies needed for training. Between April 2021 and the beginning of the study period in late 2021, Ghana received two consignments of the product totalling 240,000 units via UNFPA, which helped to improve the immediate outlook for supply in-country, however the medium- to long-term stock status at the time of the study was less certain. Stock issues have been noted as a disruption to the roll out of DMPA-SC in other contexts. [2]

Reflecting these gaps and delays in implementation, OJT providers were more likely to report fewer components in their training and dissatisfaction with many elements of their training. Furthermore, some significant differences emerged between the OJT and FT groups in their clinical knowledge of the DMPA-SC product and their recall of the critical SI steps, with OJT cohorts showing significantly poorer results on average. These differences corroborate some providers’ fears in the qualitative interviews that the quality of information reaching OJT providers was not as comprehensive as that received by the FT cohort. Interestingly, however, there were no significant differences emerging in the quality of overall injectables counselling between FT and OJT providers, likely reflecting that both populations had experience in counselling on DMPA-IM, allowing them to accommodate the new injectable option relatively easily.

The fact that confidence and skills gaps in this study emerged specifically around client training on SI is not surprising, given evidence from other studies that providers find it challenging to reassure women to overcome their initial fear of SI. [5, 7, 14, 16, 17] However, it was reassuring that a small number of entrepreneurial providers in both the FT and OJT cohort were already developing their own techniques to try and help reassure women about the SI concept – best practices that have been noted to support women’s confidence to SI in other contexts [16] and ones which could be further shared through peer-learning and supportive supervision at facility level.

It was also interesting that in this study any training (whether FT or OJT) appeared to improve provider attitudes towards the idea of a self-injectable contraceptive, when compared to untrained providers. The fact that few statistically significant differences emerged in attitude statements between FT and OJT providers is encouraging, and this likely reflects that being exposed to even some basic information about the size and ease-of-use of DMPA-SC can shift provider attitudes towards supporting self-injection. However, a few of the attitudes statements in the survey (for example, providers not trusting women to remember their re-injection dates) and qualitative examples from KIIs (for example, some providers feeling that SI is may not be suitable for women with low literacy) indicate that some trained providers may need more reassurance – either via supportive supervision or through meeting satisfied SI users - that women can and do SI safely. Providers lacking confidence that SI is possible for all clients is certainly not unique to Ghana and similar concerns have been found in other studies [8], but it could in some cases influence providers to limit offering the SI option only to certain groups of clients. Evidence from Ghana and other contexts has indicated that with a supportive provider and the right visual aids, clients of any educational status can be trained to SI. [5, 10]

It was notable that in the few facilities in this study where the OJT model was seen to be working well, this tended to be where the FT provider was an experienced provider with good training skills, who embedded regular (for example, twice per year) OJT into their work. The slight differences in length of clinical experience reported by FT and OJT providers in the provider survey suggests that facility in-charges are already bearing the providers’ seniority in mind when selecting who to attend the formal trainings, however it may help to provide non-binding guidance to facility in-charges on selecting FT providers with peer-training experience to maximize the impact of OJT. In addition, the one aspect of training OJT providers evaluated more positively than FT providers was post-training follow up, highlighting the skills-strengthening opportunities of working alongside experienced peers and directly with clients on a regular basis. This echoes a finding from studies evaluating provider acceptability of hybrid and e-learning approaches for DMPA-SC and H-IUD, where aspects such as the flexibility and convenience of being able to learn in situ were highlighted. [12, 18]

For FT and OJT providers in this study who received follow-up supportive supervision or mentorship visits, this was noted as being helpful for retaining knowledge and addressing any gaps, but almost all providers (regardless of training type) requested this support should occur more frequently. As several RRTs noted, post-training support could arguably overcome many of the challenges noted with the OJT model, for example, giving both FT and OJT providers the opportunity to practice further under observation, share best practices on SI counselling and training challenges, and address competency gaps arising from any delayed or incomplete OJT. However, at the time of the study most RRT members were not involved in any follow-up support for DMPA-SC with providers due to lack of time and funding to conduct site visits. The potential of supportive supervision to optimise alternative training approaches has also been noted in an evaluation of e-learning for DMPA-SC in Uganda and Senegal. [12]

While Ministries of Health across low- and middle-income countries are engaged in the roll out of new contraceptive products such as DMPA-SC, they will inevitably have to explore alternatives to classroom-based in-service training to increase training coverage across their workforce most cost-effectively. To date, most studies have documented the effectiveness of classroom-based training models – either within a pilot or, if at scale, cascaded down through levels of the health system in clusters or staggered geographically in clusters. [2, 10] A few studies have demonstrated the effectiveness of e-learning options at achieving comparable knowledge retention and clinical competency - although this is recommended as an adjunct to, rather than a replacement for, classroom in-service training and/or practicum. [12, 18] This study builds upon those insights to describe the challenges of ensuring consistent quality of training when implementing an alternative training model at scale and draws out recommendations to optimise such models through supportive supervision.

### Limitations

While this study provided valuable information to inform the GHS’ continued scale up of DMPA-SC for self-injection in Ghana, several limitations should be noted:

- The sample for the provider survey was designed to be representative of only sites with at least one FT provider in the four study regions (i.e. not nationally representative). The facilities prioritised for in-service training roll out were selected by District Directorates and as such may differ in unknown ways in terms of characteristics from the sites where training has not yet been rolled out. Additionally, the picture of the DMPA-SC roll-out in the four study regions may differ from other regions in Ghana.
- Identifying the geographical and sectoral distribution of the DMPA-SC trained health care workers in Ghana to establish a complete sampling frame was challenging. This was partly due to the recent national redistribution of facilities across regional and district boundaries in Ghana, plus lack of visibility over the geographic distribution of trainings conducted by private and social marketing organization partners. The most up-to-date data available on public and private sector DMPA-SC training was used to estimate as complete a sampling frame as possible.
- During quantitative data collection, many site substitutions were necessary, as many facilities originally indicated to have an FT provider in the sampling frame were found to either have lost the provider to another site, or sometimes that provider was on leave or attending a training. Where sampled sites were substituted, these were always switched for another trained site in the same district. However, due to these limitations, the resulting sampling frame (and therefore sample) may not be fully reflective of the population of trained public and private facilities in the four regions.
- When selecting study sites for the survey, the inclusion criteria (i.e. minimum thresholds of DMPA-SC training coverage) used to identify the sites in the sampling frame means that the results may not be representative of regions and districts with low coverage of DMPA-SC training. It is likely that the key outcomes (including coverage and quality of OJT training and providers’ accurate knowledge of DMPA-SC) will differ in areas where FT coverage is even lower.
- The sample size for the provider survey was only powered to detect differences in the primary knowledge outcomes between FT and OJT providers of 20 percentage points or more. Where differences in outcomes between these groups were <20 percentage points and Chi2 test results showed the difference to be non-statistically significant, this may simply indicate that the sample was not powered to detect such small differences. Any differences in knowledge and skills detected between these groups may be confounded by factors such as length of clinical experience, as providers selected for FT were on average slightly more experienced than those eligible for OJT. However, the fact that significant differences in knowledge and skills were most pronounced around specific aspects of DMPA-SC knowledge and skill, while there were few significant differences in general attitudes and injectables counselling quality between FT and OJT providers, suggests that the influence of this confounding is likely minimal.
- Purposive sampling for qualitative elements ensured that individuals with direct experience of DMPA-SC were sampled, however given the partial roll out of DMPA-SC across geographies and levels of the health system at the time of the study, this approach meant that the resulting sample could be subject to unknown bias. Every effort was made to ensure a range of characteristics in the facilities selected for qualitative data collection to minimise this.
- Provider role plays of injectables counselling and self-injection client training were conducted with enumerators trained to score providers against set criteria. It is likely that some element of Hawthorne effect influenced how providers acted out these role plays. However, providers were reassured that these scores were not a personal test and would be anonymised for analysis to minimise this bias.

## Conclusion

Cascaded in-service OJT models, such as the one GHS implemented for DMPA-SC, have great potential to increase training coverage cost-effectively. However, this study’s findings highlight the importance of ensuring that such models include adequate checks and balances to standardise the quality and timeliness of OJT, while simultaneously investing in supportive supervision structures to identify and address any residual knowledge and skills gaps among all providers, in particular on the challenging area of building client confidence to SI. The authors recommended to standardise expectations of the timeliness and content and breadth of OJT; issue optional guidance to facility in-charges to prioritise colleagues with peer-training experience for FT; improve consistency of DMPA-SC stock and associated materials to support counselling (e.g. visual aids, calendars); and improve consistency of post-training supportive supervision to address any residual knowledge and skills gap and optimise the model. Following this study, GHS implemented several of these recommendations to strengthen the OJT model in their ongoing efforts to scale up DMPA-SC across Ghana.

## Data Availability

The data collection tools, anonymised qualitative code reports (coded qualitative excerpts against relevant themes) and the anonymised provider survey dataset behind the findings and conclusions of this paper are available via https://doi.org/10.5281/zenodo.15545267 The qualitative code reports have been provided rather than full in-depth interview transcripts due to concerns about narrative descriptions and contextual references in the full qualitative transcripts making participants possibly indirectly re-identifiable. Qualitative analysis can still be re-created using the code reports provided.

## Acknowledgements

The authors would like to express our huge thanks to all colleagues from the Family Health Division at the Ghana Health Service – specifically our colleagues Dr Yaa Asante, Angela Boateng, and Rebecca Tricia Morrison – for their tireless support in implementing this study. Thanks also to CHAI colleagues who provided technical support to this study, including Forum Mistry, Emma Aldrich, and Manish Burman. Finally, sincere thanks to all the research assistants who collected our data and to the providers who graciously shared their experiences and perspectives for this research.

## Supporting information

**S1 Table. Theoretical framework outlining behavioural and contextual barriers and enablers of providers integrating DMPA-SC, including for self-injection, into routine care**

## Notes

### Competing Interest Statement

The authors have declared no competing interest.

### Funding Statement

This work was supported, in whole or in part, by the Gates Foundation [Opportunity ID: OPP1195232]. The funders had no role in study design, data collection and analysis, decision to publish, or preparation of the manuscript. The conclusions and opinions expressed in this work are those of the author(s) alone and shall not be attributed to the Foundation. Under the grant conditions of the Foundation, a Creative Commons Attribution 4.0 License has already been assigned to the Author Accepted Manuscript version that might arise from this submission.

### Author Declarations

This study was conducted in line with the principles outlined in the Declaration of Helsinki. The study team worked closely with the study regions, districts and facilities through the office of the Ghana Health Service Director General to secure appropriate approvals. The study protocol and tools were approved by GHS Ethics Review Committee in Accra, Ghana, a board that is also registered as an International Institutional Review Board with Federal Wide Assurance (IRB: IRB00009260, FWA00020025), study protocol approval number GHS-ERC: 019/08/21. All participants provided written informed consent to participate in the survey and key informant interviews.

